# Specific diagnostic criteria identify those at high risk for progression from ‘preaddiction’ to severe alcohol use disorder

**DOI:** 10.1101/2023.06.12.23291164

**Authors:** Alex P. Miller, Sally I-Chun Kuo, Emma C. Johnson, Rebecca Tillman, Sarah J. Brislin, Danielle M. Dick, Chella Kamarajan, Sivan Kinreich, John Kramer, Vivia V. McCutcheon, Martin H. Plawecki, Bernice Porjesz, Marc A. Schuckit, Jessica E. Salvatore, Howard J. Edenberg, Kathleen K. Bucholz, Jaquelyn L. Meyers, Arpana Agrawal

## Abstract

**Importance:** Both current DSM-5 diagnoses of substance use disorders (SUDs) and the recent “preaddiction” conceptual proposal (i.e., mild-to-moderate SUD) rely on criterion count-based approaches, without consideration of evidence regarding varying severity grading indexed by individual criteria.

**Objective:** To examine correlates of alcohol use disorder (AUD) across count-based severity groups (i.e., mild, moderate, mild-to-moderate, severe), identify specific diagnostic criteria indicative of greater severity, and evaluate whether the presence of specific criteria within mild-to-moderate AUD differentiates across relevant correlates and manifests in greater hazards of severe AUD development.

**Design:** Cross-sectional and longitudinal cohort study.

**Setting:** Family-based study of individuals from seven sites across the United States.

**Participants:** Cross-sectional (*N*=13,110; mean [SD] age, 37.8 [14.2] years) and longitudinal cohorts (*N*=2,818; mean baseline [SD] age, 16.1 [3.2] years) from the Collaborative Study on the Genetics of Alcoholism (COGA).

**Exposure:** N/A

**Main Outcomes and Measures:** Sociodemographic, alcohol-related, psychiatric comorbidity (major depressive disorder, antisocial personality disorder, and other SUDs), brain electroencephalography (EEG), and AUD polygenic score measures as correlates of DSM-5 AUD levels (i.e., mild, moderate, severe) and criterion severity-defined “preaddiction” (i.e., low-risk *vs.* high-risk mild-to-moderate) AUD diagnostic groups.

**Results:** Associations with alcohol-related, psychiatric, EEG, and AUD polygenic score measures reinforced the role of increasing criterion counts as indexing severity. Yet even within those meeting criteria for mild-to-moderate AUD (2-5 criteria), the presence of specific “high-risk” criteria (e.g., withdrawal) identified a group reporting heavier drinking and greater psychiatric comorbidity even after accounting for criterion count differences. In longitudinal analyses, prior mild-to-moderate AUD characterized by endorsement of at least one “high-risk” criterion outperformed other adolescent and young adult correlates of AUD progression (i.e., comorbid psychiatric diagnoses, alcohol involvement milestones) and was associated with more accelerated progression to severe AUD (adjusted hazard ratio [aHR], 11.62; 95% CI, 7.54-17.92) compared to prior mild-to-moderate AUD without endorsement of “high-risk” criteria (aHR, 5.64; 95% CI, 3.28-9.70), independent of criterion count.

**Conclusions and Relevance:** Current count-based AUD diagnostic approaches and the “preaddiction” concept both ignore heterogeneity among criteria. Estimating addiction vulnerability by emphasizing specific “high-risk” criteria may improve our understanding of its development and focus attention on those at greatest risk.

**Key Points:** *Question:* Does emphasis on specific alcohol use disorder (AUD) criteria improve identification of individuals at risk for developing more severe AUD?

*Findings:* Individuals meeting criteria for mild-to-moderate AUD are two-fold more likely to progress to severe AUD if they endorse criteria for drinking despite physical/psychological problems, giving up important activities, spending a great deal of time drinking, failure to fulfill major role obligations, withdrawal, and craving, even after accounting for total criterion count.

*Meaning:* Emphasis on especially severe criteria as indicators of addiction vulnerability in current diagnostic approaches may increase detection of individuals with greater likelihood for disorder progression.

## Introduction

Alcohol use disorder (AUD, as defined by the DSM-5^1^) is conceptualized as a syndrome of sustained problematic alcohol use and clinically significant impairment. A diagnosis is based on endorsement of two or more of 11 criteria assessing behavioral and physical manifestations of heavy alcohol use that occur in a 12-month period.^1^ Recent estimates in the U.S. indicate that 11% of adults meet criteria for a past-year AUD,^2^ and the lifetime prevalence rate of AUD has been estimated to be as high as 30%.^3^ This level of disordered alcohol use in the general population results in significant social, economic, and public health costs.^4–6^

Identifying individuals at greater risk of developing chronic and severe forms of AUD is a priority. In the DSM-5, AUD is diagnosed on a continuum of severity based on the number of criteria endorsed (2-3=mild, 4-5=moderate, ≥6=severe)^1^. Several studies document increased comorbid burden and reduced likelihood of recovery as a function of increasing criterion count.^7–9^ In a recent proposal, McLellan et al.^10^ suggest that individuals endorsing 2-5 DSM-5 substance use disorder (SUD) diagnostic criteria (mild-to-moderate SUD) may be conceptualized as those with early-stage SUDs, termed “preaddiction” by the authors (similar to prediabetes). The authors further posit that those with “preaddiction” are at elevated risk for progression to “addiction”, a term they use to refer to severe SUD (≥6 SUD criteria).

Criterion count-based severity indices are limited by their equal weighting of all diagnostic criteria, suggesting criteria are interchangeable. However, extensive cross-national psychometric evidence shows that certain criteria might be superior indicators of severity and risk likelihood (e.g., ^11–13^). For AUD specifically, research has demonstrated that certain criteria (withdrawal, giving up important activities, craving) are more likely to be endorsed by those at above average risk for AUD.^14, 15^ Thus, criteria heterogeneity is an important factor to consider in the advancement of personalized treatment approaches.^16^

In light of evidence suggesting that diagnostic categorization based solely on criterion count may be suboptimal for identifying individuals at higher risk for developing severe AUD, we investigate the impact of criteria heterogeneity within criterion count-based AUD diagnoses (i.e., mild *vs.* moderate *vs.* severe; and mild-to-moderate *vs.* severe [“preaddiction” *vs.* “addiction” as proposed by McLellan et al.^10^]) using multimodal indices of AUD severity. The current study sought to (1) validate criterion count-based severity, using both DSM-5 and McLellan et al.^10^ categorizations of AUD, in a sample of 13,110 individuals from a family-based study with deep and repeated phenotyping of SUDs, comorbid psychiatric disorders, and related traits (including polygenic liability and electrophysiological markers); (2) utilize item response theory (IRT) modeling to identify criteria indicative of greater severity; and (3) evaluate whether the presence of certain high-risk criteria, identified through IRT modeling, index greater hazards of developing severe AUD in a related cohort of 2,818 adolescents and young adults.

## Methods

### Participants

The Collaborative Study on the Genetics of Alcoholism (COGA) is a family-based study designed to examine the genetic substrates of AUD and its development across the lifespan.^17^ AUD probands were recruited primarily from treatment facilities across seven U.S. collection sites. Comparison families were also included, selected from a variety of community sources. The Institutional Review Boards at all seven sites approved this study, and written consent was obtained from all participants. We follow the STROBE reporting guidelines for cohort studies. Data were restricted to alcohol-exposed individuals (i.e., endorsing lifetime use of any alcohol) from two COGA sub-samples: (1) a cross-sectional cohort of 13,110 individuals from 2,234 families assessed from 1991-2005 (mean [SD] age, 37.8 [14.2] years, 52.8% female, 22.8% African American), and (2) a cohort of 2,818 offspring of individuals in the cross-sectional cohort, born after 1981, comprising the longitudinal component of COGA (mean baseline [SD] age, 16.1 [3.2] years, 52.5% female, 26.4% African American).^18^

### Measures

#### Cross-Sectional Cohort

The cross-sectional cohort was used to (1) examine correlates of mild, moderate, mild-to-moderate, and severe AUD; (2) identify AUD criteria indicative of heightened risk using item response theory (IRT) analysis; and (3) evaluate whether individuals with mild-to-moderate AUD who endorsed high-risk criteria differed from those who did not, and from those with severe AUD, across relevant factors, including alcohol-related, comorbid psychiatric, and EEG-derived traits and polygenic indices.

AUD criteria, diagnoses, and several correlates were derived from the Semi-Structured Interview for the Genetics of Alcoholism (SSAGA).^19, 20^ Sociodemographic variables included sex, race/ethnicity, current income, educational attainment, and relationship status. Psychiatric lifetime DSM-IV^21^ diagnoses included major depressive disorder (MDD), antisocial personality disorder (ASPD), and other SUD diagnoses (endorsing ≥2 DSM-5 criteria for cannabis, cocaine, opiate, stimulant, sedative, or ‘other’ use disorder, or DSM-IV nicotine dependence). In addition, several alcohol-related measures were included as correlates in cross-sectional analyses: (1) lifetime endorsement of drinking every day for a week or more; (2) largest number of drinks consumed each day during this period; (3) lifetime endorsement of ‘blackouts’; (4) age of first intoxication; (5) age of regular drinking onset (i.e., drinking once per month for six months or more); (6) lifetime maximum number of drinks ever consumed in a single 24-hour period; and (7) lifetime endorsement of seeking professional help or engaging in treatment for drinking problems.

COGA includes electroencephalography (EEG)-derived event-related oscillatory response measures (EROs).^22^ Prior studies have found the P300 component during the standard visual oddball paradigm to be associated with family history of AUD.^22^ Subgroup differences for parietal delta (1-3 Hz) and frontal theta (3-7 Hz) band EROs (300-700 ms window) and parietal P300 amplitude were also included as correlates of AUD severity in cross-sectional analyses. Polygenic scores (PGS) for AUD diagnostic status (i.e., case/control), derived from a meta-analysis of large-scale genome-wide association study (GWAS) summary statistics,^23–26^ were calculated for genotyped individuals of European (EA; *n*=5,396) and African (AA; *n*=1,774) ancestry separately using PRS-CS-auto^27^ and PRS-CSx,^28^ respectively (**Supplement eMethods**).

#### Longitudinal Cohort

The longitudinal sample included participants aged 11-26 years at baseline interview who were followed approximately every two years (mean [SD] number of timepoints, 3.2 [1.8]). A SSAGA was administered at each biennial assessment. Given the longitudinal design, this sample was used to examine whether prior mild-to-moderate AUD diagnoses, with and without endorsement of high-risk criteria, were associated with increased hazards of progression to severe AUD. Effects of prior single lifetime criterion endorsement, mild AUD (2-3 criteria), and moderate AUD (4-5 criteria) as well as other well-studied correlates, including alcohol involvement milestones (i.e., age of first drink, regular drinking, and intoxication), and MDD, ASPD, and SUD diagnoses were likewise tested to examine whether these exerted comparable effects to prior diagnosis of mild-to-moderate AUD.

### Statistical Analyses

#### Cross-Sectional Cohort

First, means and prevalence of sociodemographic, alcohol-related, and comorbid psychiatric measures were evaluated across AUD severity categories (mild, moderate, mild-to-moderate, and severe). Statistical comparisons of mild *vs.* moderate AUD and moderate *vs.* severe AUD were conducted across alcohol-related and comorbid psychiatric measures using Wilcoxon rank sum and Fisher’s exact tests to evaluate the validity of combining mild and moderate to represent mild-to-moderate AUD (“preaddiction” as defined by McLellan et al.^10^). Second, a one-parameter logistic IRT model assuming unidimensional structure for the 11 AUD criteria was applied to estimate criteria difficulty (severity) using the *mirt* package (v1.37.1)^29^ in R^30^ (**Supplement eMethods**). Criteria were then rank ordered by severity and those with severity parameter values >2 (i.e., 50% endorsement probability by individuals 2+ *SD* above mean AUD latent severity) were considered “high-risk” (**Figure 1; Supplement eTable 1**). Third, individuals with mild-to-moderate AUD either endorsing no “high-risk” criteria or at least one “high-risk” criterion were classified into “low-risk” (*n*=2,486) and “high-risk” (*n*=993) groups, respectively. Low-risk mild-to-moderate, high-risk mild-to-moderate, and severe AUD groups were then compared across measured alcohol-related, psychiatric, and EEG correlates using mixed effects logistic and linear regression models fitted using the *lme4* package (v1.1-30)^31^ in R controlling for sex, age, race/ethnicity, and criterion count. Because COGA includes related individuals nested within family, mixed effects models adjusted for familial clustering. Similar mixed effects models, conducted separately by ancestry, were used to examine associations between AUD PGSs and diagnostic groups (**Supplement eMethods**).

**Figure 1.**
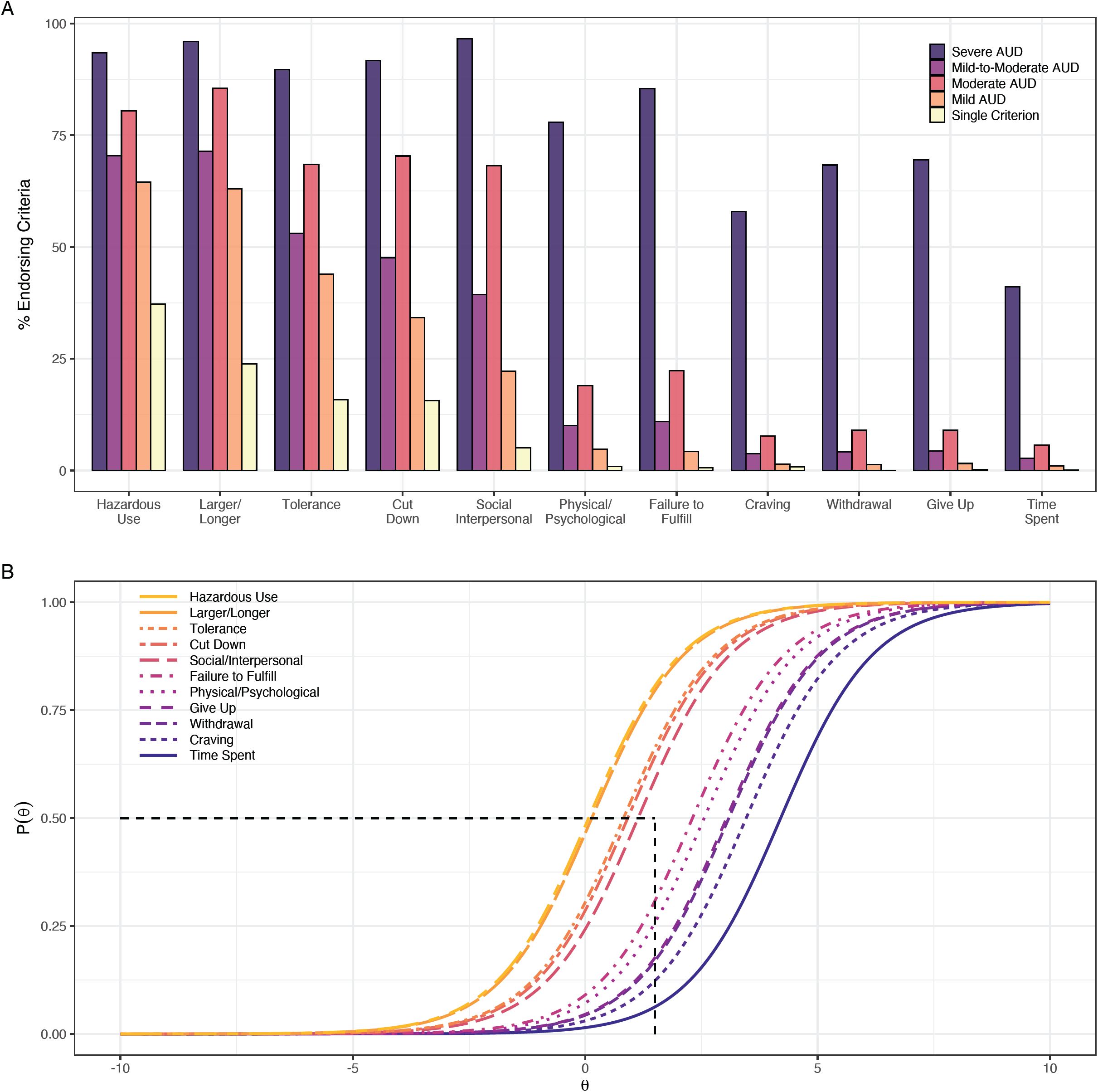
Percent endorsement by diagnostic group and IRT response curves for 11 lifetime AUD criteria in cross-sectional COGA cohort (*N*=13,110). Panel B: The probability of endorsement of each AUD criteria (P(Θ); *Y*-axis) is plotted as a function of increasing severity of the underlying AUD latent trait (Θ; *X*-axis). Horizontal dashed line represents 50% probability of endorsing criteria, vertical dashed line represents 2 *SD* above mean of AUD latent severity. Criteria with difficulty parameters of 2 or above (i.e., to the right of intersection of vertical and horizontal lines: failure to fulfill, physical/psychological, give up, withdrawal, craving, and time spent) were identified as “high-risk” diagnostic criteria. Hazardous Use = Recurrent alcohol use (3+ times) in situations in which it is physically hazardous; Larger/longer = Drinking in larger amounts or over longer periods than intended; Tolerance = Need for markedly increased amounts of alcohol to achieve intoxication or desired effect or a markedly dimin-ished effect with continued use of the same amount of alcohol; Cut Down = Persistent desire or three or more unsuccessful efforts to stop, cut down, or control drinking; Social Interpersonal = Continued alcohol use despite having persistent or recurrent (3+ times) social or interpersonal problems caused or exacerbated by the effects of alcohol; Failure to Fulfill = Recurrent use of alcohol resulting in a failure to fulfill major role obligations at work, school, or home; Physical/Psychological = Continued drinking despite knowledge of having a persistent or recurrent physical or psychological problem that is likely to be caused or exacerbated by drinking; Give Up = Important social, occupational, or recreational activities given up or reduced because of drinking; Withdrawal = The characteristic withdrawal syndrome for alcohol; or drinking (or using a closely related substance) to relieve or avoid withdrawal symp-toms; Craving = Craving, or a strong desire or urge to use alcohol; Time Spent = A great deal of time spent in activities necessary to obtain, to use, or to recover from the effects of drinking

#### Longitudinal Cohort

Hazards of progression to severe AUD (≥6 criteria) were estimated for those who, at any previous timepoint, had endorsed a single criterion or met criteria for mild, moderate, or mild-to-moderate AUD via Cox proportional hazards survival analyses conducted using the *survival* (v3.4-0)^32^ and *adjustedCurves* (v0.9.1)^33^ packages in R. Low-risk (*n*=699) and high-risk (*n*=317) mild-to-moderate AUD subgroups were defined by a prior mild-to-moderate AUD diagnosis and differentiated by endorsement of one or more high-risk criteria identified in the cross-sectional IRT analysis. Additional survival analyses were conducted examining hazards of progression to severe AUD based on alcohol involvement milestones, MDD, ASPD, and SUD diagnoses. Covariates for all models included sex, race/ethnicity, and criterion count, and family grouping variable was used to estimate robust standard errors to account for familial clustering.^34^ Violations of the proportional hazards assumption for predictor variables were tested using Schoenfeld residuals and resolved by including an interaction term with age at onset of severe AUD.

## Results

### Differences across criterion count-based AUD groups

As expected, increasing criterion count (i.e., single criterion, mild, moderate, and severe AUD) was associated with increasing levels of heavy alcohol use and greater psychiatric comorbidity (**Table 1**). For instance, 48.9% and 82.4% of those with mild-to-moderate and severe AUD, respectively, reported experiencing blackouts. Likewise, 80% of those with severe AUD met criteria for a comorbid SUD *vs.* half of those with mild-to-moderate AUD. Count-based severity was also reflected in reduced P300 amplitude and theta and delta EROs in those with severe *vs.* mild-to-moderate AUD. AUD criterion count and diagnostic status (AUD *vs.* no AUD) were associated with increased genetic liability for AUD across ancestral subsamples (**Supplement eTable 3**). AUD polygenic scores (PGSs) also differentiated between severe and no AUD in the EA subsample, (odds ratio [OR], 1.23; 95% CI, 1.12-1.35), and between severe and mild AUD in the AA subsample (OR, 1.27; 95% CI, 1.07-1.51). Those with mild AUD differed from those with moderate AUD on alcohol-related and psychiatric variables, but overall, effect size differences between these two severity groups (together comprising the McLellan et al.^10^ definition of “preaddiction”) were less pronounced than those between moderate and severe AUD, or between mild-to-moderate and severe AUD.

**Table 1.**
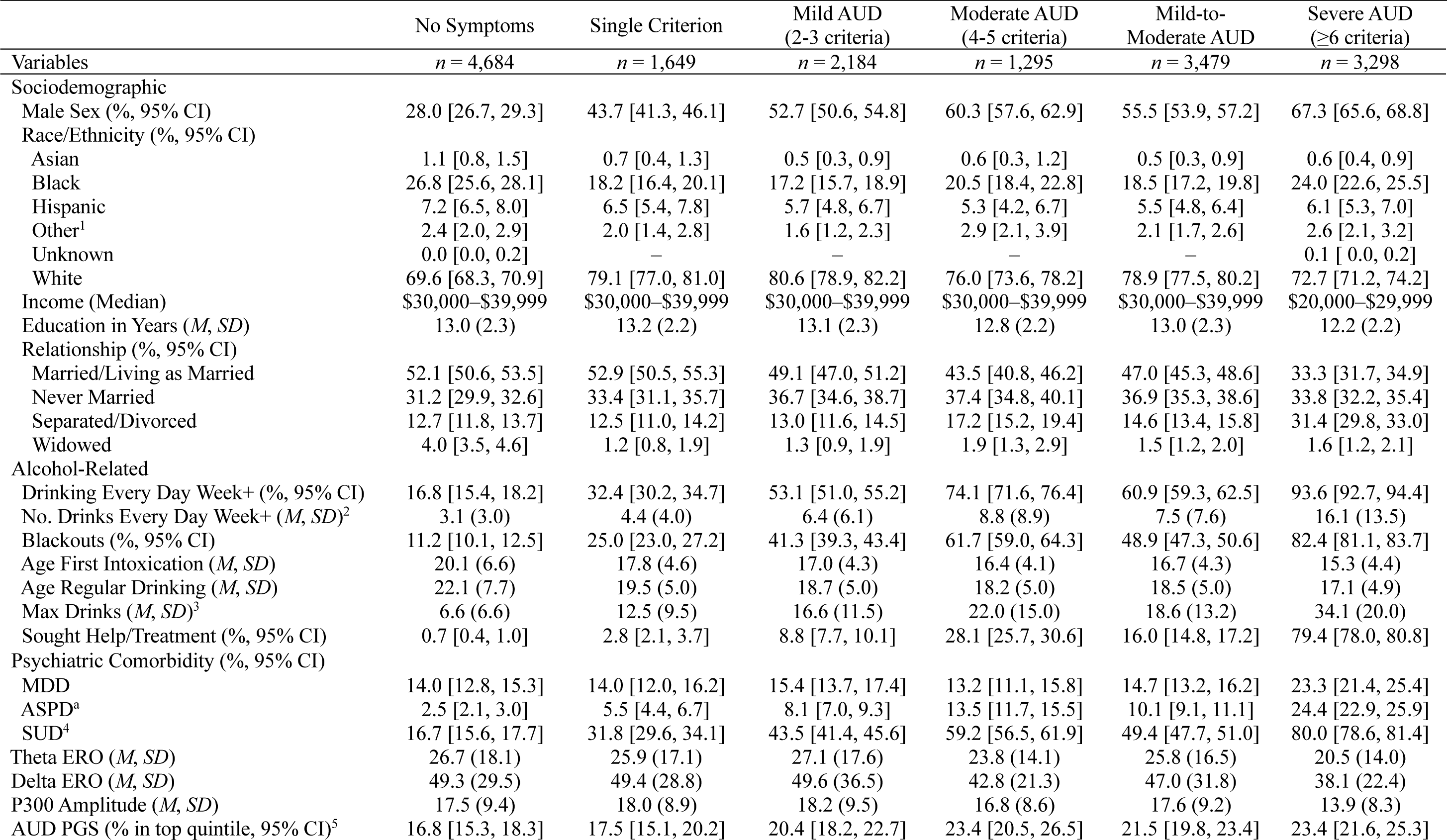
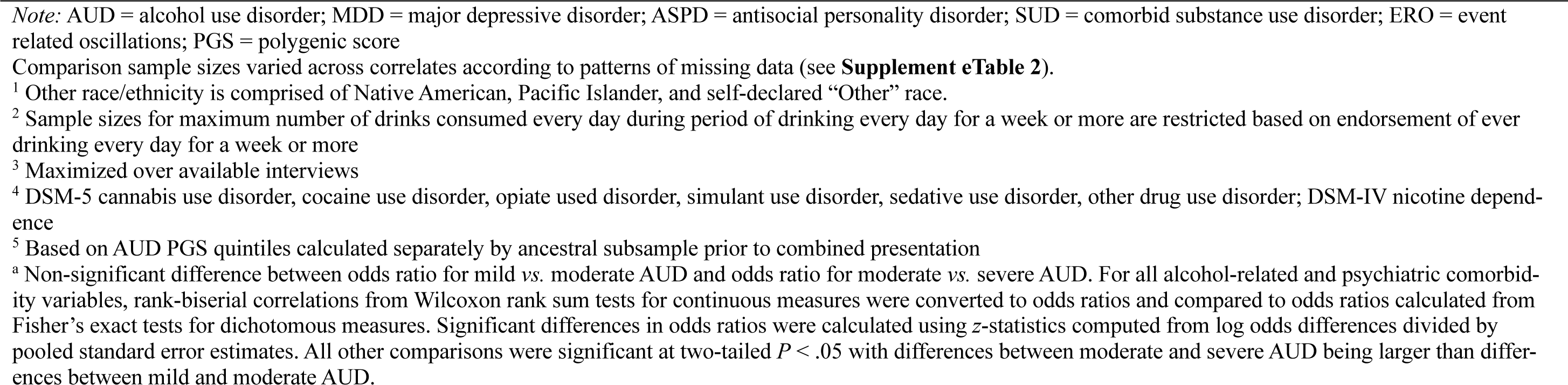
Cross-sectional COGA cohort (*N*=13,110) descriptive statistics for sociodemographic, alcohol-related, psychiatric comorbidity, electroencephalography, and AUD polygenic score correlates organized by diagnostic groups

| | No Symptoms | Single Criterion | Mild AUD<br>(2-3 criteria) | Moderate AUD<br>(4-5 criteria) | Mild-to-Moderate AUD | Severe AUD<br>($\geq 6$ criteria) |
| --- | --- | --- | --- | --- | --- | --- |
| Variables | $n = 4,684$ | $n = 1,649$ | $n = 2,184$ | $n = 1,295$ | $n = 3,479$ | $n = 3,298$ |
| **Sociodemographic** |  |  |  |  |  |  |
| Male Sex (%; 95% CI) | 28.0 [26.7, 29.3] | 43.7 [41.3, 46.1] | 52.7 [50.6, 54.8] | 60.3 [57.6, 62.9] | 55.5 [53.9, 57.2] | 67.3 [65.6, 68.8] |
| Race/Ethnicity (%; 95% CI) |  |  |  |  |  |  |
| Asian | 1.1 [0.8, 1.5] | 0.7 [0.4, 1.3] | 0.5 [0.3, 0.9] | 0.6 [0.3, 1.2] | 0.5 [0.3, 0.9] | 0.6 [0.4, 0.9] |
| Black | 26.8 [25.6, 28.1] | 18.2 [16.4, 20.1] | 17.2 [15.7, 18.9] | 20.5 [18.4, 22.8] | 18.5 [17.2, 19.8] | 24.0 [22.6, 25.5] |
| Hispanic | 7.2 [6.5, 8.0] | 6.5 [5.4, 7.8] | 5.7 [4.8, 6.7] | 5.3 [4.2, 6.7] | 5.5 [4.8, 6.4] | 6.1 [5.3, 7.0] |
| Other<sup>1</sup> | 2.4 [2.0, 2.9] | 2.0 [1.4, 2.8] | 1.6 [1.2, 2.3] | 2.9 [2.1, 3.9] | 2.1 [1.7, 2.6] | 2.6 [2.1, 3.2] |
| Unknown | 0.0 [0.0, 0.2] | — | — | — | — | 0.1 [0.0, 0.2] |
| White | 69.6 [68.3, 70.9] | 79.1 [77.0, 81.0] | 80.6 [78.9, 82.2] | 76.0 [73.6, 78.2] | 78.9 [77.5, 80.2] | 72.7 [71.2, 74.2] |
| Income (Median) | \$30,000–\$39,999 | \$30,000–\$39,999 | \$30,000–\$39,999 | \$30,000–\$39,999 | \$30,000–\$39,999 | \$20,000–\$29,999 |
| Education in Years ($M$, $SD$) | 13.0 (2.3) | 13.2 (2.2) | 13.1 (2.3) | 12.8 (2.2) | 13.0 (2.3) | 12.2 (2.2) |
| Relationship (%; 95% CI) |  |  |  |  |  |  |
| Married/Living as Married | 52.1 [50.6, 53.5] | 52.9 [50.5, 55.3] | 49.1 [47.0, 51.2] | 43.5 [40.8, 46.2] | 47.0 [45.3, 48.6] | 33.3 [31.7, 34.9] |
| Never Married | 31.2 [29.9, 32.6] | 33.4 [31.1, 35.7] | 36.7 [34.6, 38.7] | 37.4 [34.8, 40.1] | 36.9 [35.3, 38.6] | 33.8 [32.2, 35.4] |
| Separated/Divorced | 12.7 [11.8, 13.7] | 12.5 [11.0, 14.2] | 13.0 [11.6, 14.5] | 17.2 [15.2, 19.4] | 14.6 [13.4, 15.8] | 31.4 [29.8, 33.0] |
| Widowed | 4.0 [3.5, 4.6] | 1.2 [0.8, 1.9] | 1.3 [0.9, 1.9] | 1.9 [1.3, 2.9] | 1.5 [1.2, 2.0] | 1.6 [1.2, 2.1] |
| **Alcohol-Related** |  |  |  |  |  |  |
| Drinking Every Day Week+ (%; 95% CI) | 16.8 [15.4, 18.2] | 32.4 [30.2, 34.7] | 53.1 [51.0, 55.2] | 74.1 [71.6, 76.4] | 60.9 [59.3, 62.5] | 93.6 [92.7, 94.4] |
| No. Drinks Every Day Week+ ($M$, $SD$)<sup>2</sup> | 3.1 (3.0) | 4.4 (4.0) | 6.4 (6.1) | 8.8 (8.9) | 7.5 (7.6) | 16.1 (13.5) |
| Blackouts (%; 95% CI) | 11.2 [10.1, 12.5] | 25.0 [23.0, 27.2] | 41.3 [39.3, 43.4] | 61.7 [59.0, 64.3] | 48.9 [47.3, 50.6] | 82.4 [81.1, 83.7] |
| Age First Intoxication ($M$, $SD$) | 20.1 (6.6) | 17.8 (4.6) | 17.0 (4.3) | 16.4 (4.1) | 16.7 (4.3) | 15.3 (4.4) |
| Age Regular Drinking ($M$, $SD$) | 22.1 (7.7) | 19.5 (5.0) | 18.7 (5.0) | 18.2 (5.0) | 18.5 (5.0) | 17.1 (4.9) |
| Max Drinks ($M$, $SD$)<sup>3</sup> | 6.6 (6.6) | 12.5 (9.5) | 16.6 (11.5) | 22.0 (15.0) | 18.6 (13.2) | 34.1 (20.0) |
| Sought Help/Treatment (%; 95% CI) | 0.7 [0.4, 1.0] | 2.8 [2.1, 3.7] | 8.8 [7.7, 10.1] | 28.1 [25.7, 30.6] | 16.0 [14.8, 17.2] | 79.4 [78.0, 80.8] |
| **Psychiatric Comorbidity (%; 95% CI)** |  |  |  |  |  |  |
| MDD | 14.0 [12.8, 15.3] | 14.0 [12.0, 16.2] | 15.4 [13.7, 17.4] | 13.2 [11.1, 15.8] | 14.7 [13.2, 16.2] | 23.3 [21.4, 25.4] |
| ASPD<sup>a</sup> | 2.5 [2.1, 3.0] | 5.5 [4.4, 6.7] | 8.1 [7.0, 9.3] | 13.5 [11.7, 15.5] | 10.1 [9.1, 11.1] | 24.4 [22.9, 25.9] |
| SUD<sup>4</sup> | 16.7 [15.6, 17.7] | 31.8 [29.6, 34.1] | 43.5 [41.4, 45.6] | 59.2 [56.5, 61.9] | 49.4 [47.7, 51.0] | 80.0 [78.6, 81.4] |
| Theta ERO ($M$, $SD$) | 26.7 (18.1) | 25.9 (17.1) | 27.1 (17.6) | 23.8 (14.1) | 25.8 (16.5) | 20.5 (14.0) |
| Delta ERO ($M$, $SD$) | 49.3 (29.5) | 49.4 (28.8) | 49.6 (36.5) | 42.8 (21.3) | 47.0 (31.8) | 38.1 (22.4) |
| P300 Amplitude ($M$, $SD$) | 17.5 (9.4) | 18.0 (8.9) | 18.2 (9.5) | 16.8 (8.6) | 17.6 (9.2) | 13.9 (8.3) |
| AUD PGS (% in top quintile; 95% CI)<sup>5</sup> | 16.8 [15.3, 18.3] | 17.5 [15.1, 20.2] | 20.4 [18.2, 22.7] | 23.4 [20.5, 26.5] | 21.5 [19.8, 23.4] | 23.4 [21.6, 25.3] |

*Note:* AUD = alcohol use disorder; MDD = major depressive disorder; ASPD = antisocial personality disorder; SUD = comorbid substance use disorder; ERO = event related oscillations; PGS = polygenic score
Comparison sample sizes varied across correlates according to patterns of missing data (see **Supplement eTable 2**).
<sup>1</sup> Other race/ethnicity is comprised of Native American, Pacific Islander, and self-declared “Other” race.
<sup>2</sup> Sample sizes for maximum number of drinks consumed every day during period of drinking every day for a week or more are restricted based on endorsement of ever drinking every day for a week or more
<sup>3</sup> Maximized over available interviews
<sup>4</sup> DSM-5 cannabis use disorder, cocaine use disorder, opiate use disorder, stimulant use disorder, sedative use disorder, other drug use disorder; DSM-IV nicotine dependence
<sup>5</sup> Based on AUD PGS quintiles calculated separately by ancestral subsample prior to combined presentation
<sup>a</sup> Non-significant difference between odds ratio for mild vs. moderate AUD and odds ratio for moderate vs. severe AUD. For all alcohol-related and psychiatric comorbidity variables, rank-biserial correlations from Wilcoxon rank sum tests for continuous measures were converted to odds ratios and compared to odds ratios calculated from Fisher’s exact tests for dichotomous measures. Significant differences in odds ratios were calculated using $z$ -statistics computed from log odds differences divided by pooled standard error estimates. All other comparisons were significant at two-tailed $P < .05$ with differences between moderate and severe AUD being larger than differences between mild and moderate AUD.

### High- and low-risk criteria

IRT analysis of the 11 DSM-5 criteria in the cross-sectional sample revealed that drinking despite physical/psychological problems, giving up important activities, spending a great deal of time drinking, failure to fulfill major role obligations due to alcohol use, withdrawal, and craving represented greater “difficulty” of endorsement, indicating greater risk (**Figure 1; Supplement eTable 1**). Endorsement of these six high difficulty criteria differed considerably across criterion count-based severity groups (single criteria, mild, moderate, and severe AUD). For instance, withdrawal was endorsed by 4.2% of those with mild-to-moderate AUD and, in contrast, by 68.3% of those with severe AUD. Similarly, those with mild AUD were less likely to endorse high-risk criteria (e.g., 1.3% endorsing withdrawal) compared to those with moderate AUD (e.g., 9.0% endorsing withdrawal).

### Distinctions between individuals with high- and low-risk mild-to-moderate AUD, and severe AUD

After grouping individuals based on endorsement of high-risk criteria identified in the IRT analysis, those with high-risk mild-to-moderate AUD were more likely to endorse a greater number of criteria than those with low-risk mild-to-moderate AUD (e.g., 42.5% *vs.* 6.8% endorsed five criteria). Even after accounting for the number of criteria endorsed, those with mild-to-moderate AUD who endorsed high-risk criteria were more likely to consume a greater number of drinks during heavy drinking episodes and periods of frequent drinking, endorse seeking help or treatment, meet criteria for other SUDs, MDD, and ASPD, and have lower theta EROs when compared with those with low-risk mild-to-moderate AUD (**Table 2; Supplement eTable 5**). AUD PGSs also distinguished between low-risk mild-to-moderate and severe AUD in the AA subsample (OR, 1.30; 95% CI, 1.09-1.56). Notably, the high-risk mild-to-moderate group did not statistically differ from the severe AUD group on theta EROs or P300 amplitude after accounting for criterion count differences.

**Table 2.**
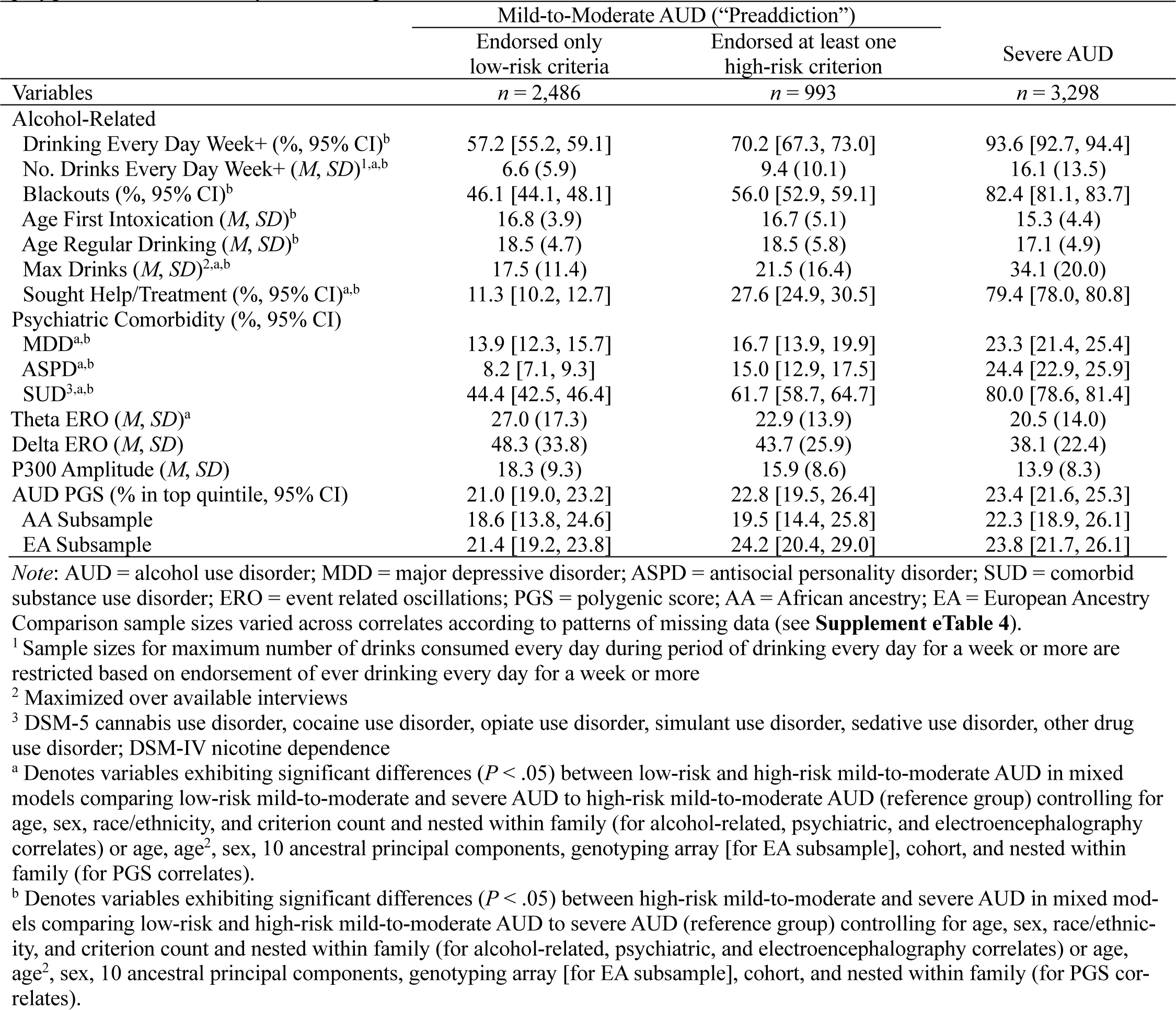
Cross-sectional COGA cohort comparisons of alcohol-related, psychiatric comorbidity, electroencephalography, and AUD polygenic score correlates by low- and high-risk mild-to-moderate and severe AUD

### Hazards of progression to severe AUD

An overwhelming majority of individuals who met criteria for severe AUD (77.8%) had a prior history of mild-to-moderate AUD (9.5% endorsed a single prior criterion, 12.7% endorsed no prior criteria). Consistent with cross-sectional findings, those with high-risk mild-to-moderate AUD were more likely (33.4%) to transition to severe AUD than those with low-risk mild-to-moderate AUD (12.9% transitioned to severe AUD; **Table 3; Supplement eResults**). The hazards of transitioning from high-risk mild-to-moderate AUD to severe AUD (adjusted hazard ratio [aHR], 11.62; 95% CI, 7.54-17.92) was more than double that of transitioning to severe AUD from low-risk mild-to moderate AUD after accounting for criterion count (aHR, 5.64; 95% CI, 3.28-9.70; between-groups-aHR, 2.06; 95% CI, 1.47-2.88; **Figure 2**). Earlier age of first drink, regular drinking, and first intoxication and comorbid ASPD, MDD, and SUDs were significantly associated with progression to severe AUD; however, hazards for these characteristics were considerably lower than hazards for belonging to the high-risk mild-to-moderate AUD group. In multivariate models, high-risk mild-to-moderate was the strongest predictor of progression to severe AUD (aHR, 4.25; 95% CI, 2.57-7.04).

**Figure 2.**
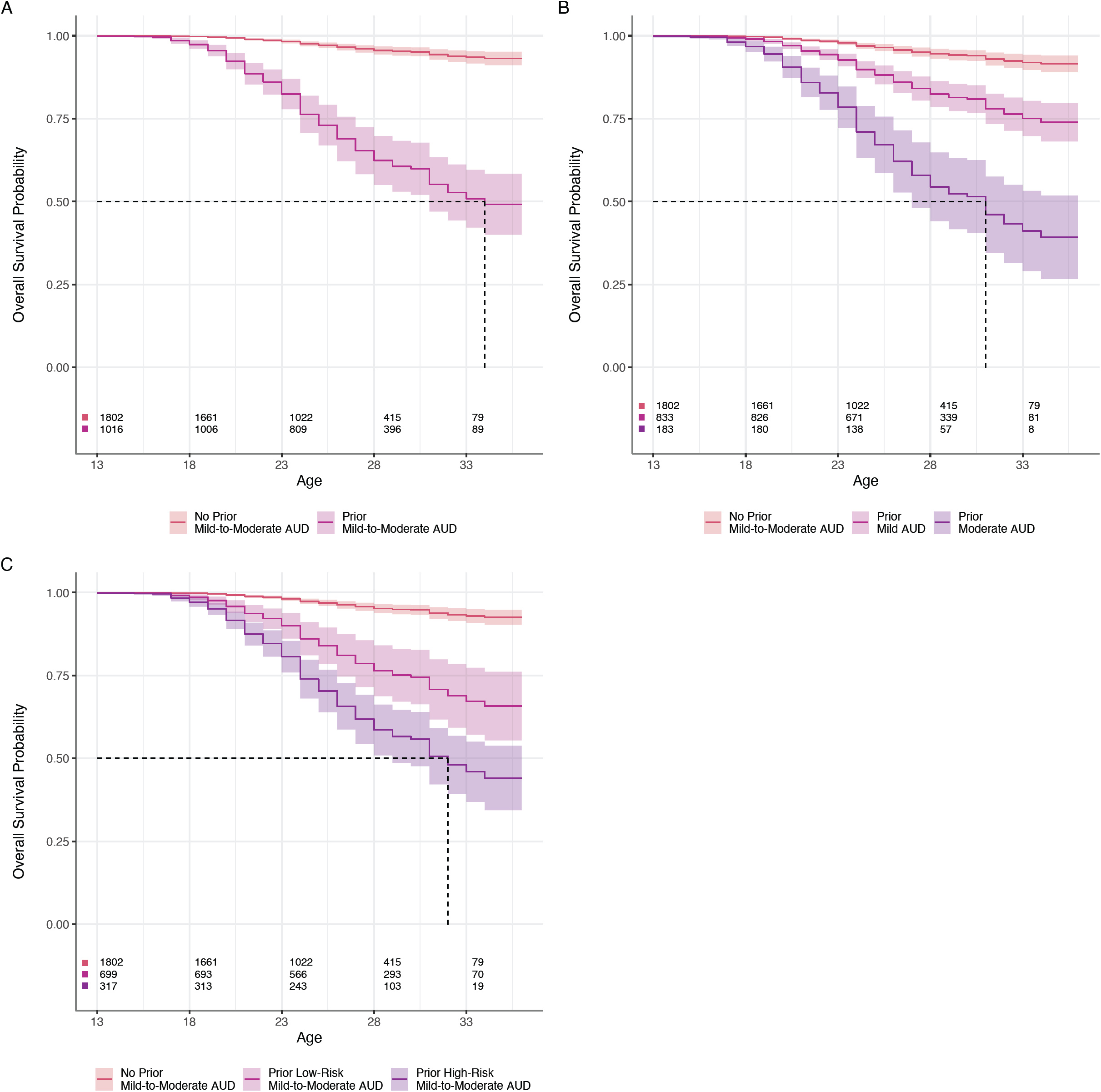
Cumulative survival curves with 95% confidence intervals for associations between prior mild-to-moderate AUD and progression to severe AUD in longitudinal COGA cohort (*N*=2,818). All survival curves include no prior mild-to-moderate AUD as comparison and are adjusted for sex, race/ethnicity, and mild-to-moderate AUD criterion count. Panel A: Survival curves additionally adjusted for endorsement of prior mild-to-moderate AUD. Panel B: Survival curves additionally adjusted for mild vs. moderate AUD. Panel C: Survival curves additionally adjusted for low-risk vs. high-risk mild-to-moderate AUD. Dashed lines represent point estimates of median survival ages (not accounting for 95% CIs): A. Prior mild-to-moderate AUD = 34 years; B. Mild AUD = undefined, moderate AUD = 31 years; C. Low-risk mild-to-moderate AUD = undefined, high-risk mild-to-moderate AUD = 32 years

**Table 3.**
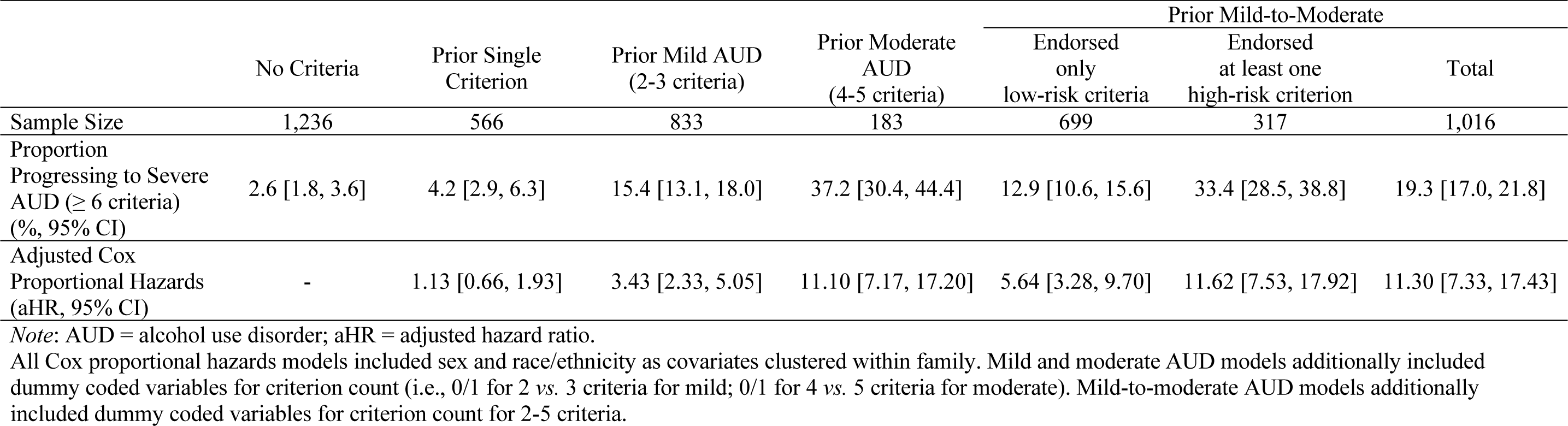
Longitudinal COGA cohort (*N*=2,818) results from Cox proportional hazards models for progression to severe AUD

## Discussion

The clinical definition of AUD is based upon a count of criteria, as established by DSM-5.^1^ However, equal weighting of criteria contributes to heterogeneity within diagnostic categories, particularly at intermediate count-based diagnostic levels.^35^ We sought to examine whether the endorsement of certain criteria elevated risk for severe AUD within the DSM-5 diagnostic scheme, in part motivated by the recent “preaddiction” concept^10^.

Results from our IRT analyses revealed six “high-risk” criteria of the 11 DSM-5 criteria that reflect greater severity. While increasing criterion counts reflected increasingly severe drinking, psychiatric, EEG and polygenic risk in the cross-sectional cohort, there were significant differences among those who endorsed high-*vs.* low-risk criteria. Moreover, individuals with mild-to-moderate AUD, or “preaddiction”,^10^ who endorsed one of these high-risk criteria were statistically indistinguishable from those with severe AUD with respect to theta EROs and P300 amplitude (after accounting for differences in criterion count). In the longitudinal cohort, individuals with moderate AUD or high-risk mild-to-moderate AUD had the greatest hazards of progression to severe AUD across late adolescence and young adulthood. Overall, these analyses suggest that the presence of specific criteria are a superior indicator of risk for progression to severe AUD compared to criterion count alone. This difference is especially pronounced for the mild-to-moderate AUD group (i.e., “preaddiction” as proposed by McLellan et al.^10^).

Many of the six empirically identified “high-risk” criteria have previously demonstrated stronger associations with comorbid psychopathology and greater severity of and discriminatory capabilities for AUD than other criteria.^14, 36–38^ However, these high-risk criteria do not currently comprise a coherent DSM-5 AUD subtype.^39^ For example, though DSM-5 characterizes both withdrawal and tolerance as physiological components of AUD, withdrawal, but not tolerance, was identified as high-risk. Interestingly, several of the high-risk criteria were markers of preoccupation (e.g., craving, time spent, giving up important activities) and impairment in several domains (e.g., role obligations, recurrent physical/psychological problems, withdrawal). These criteria were endorsed far more frequently by those with severe AUD and map onto two of the key stages (preoccupation/anticipation and withdrawal/negative affect) of the neurobiological model of addiction, which was developed to characterize severe forms of SUDs.^40^

Importantly, we were able to study neurobiological markers of AUD in the COGA sample. Unlike heavy drinking or psychiatric comorbidity, which may arise as a consequence of problematic drinking patterns, polygenic scores (PGSs) and EEG measures provide a glimpse into potential neurobiological vulnerabilities influencing distinctions across severity groups. Overall, EEG and AUD PGS results generally echoed other findings. Specifically, theta EROs distinguished between those with low-risk and high-risk mild-to-moderate AUD and demonstrated similarly blunted neurophysiological responses in both high-risk mild-to-moderate and severe AUD groups in the cross-sectional sample. AUD PGSs primarily exhibited associations with criterion count and overall diagnostic status (AUD *vs.* no AUD) across both ancestry subsamples. This is not unexpected as the genetic liability indexed by these PGSs is derived from GWAS of AUD diagnoses with a preponderance of electronic health record-derived diagnostic codes,^23–26^ amplifying the need for criterion-focused discovery efforts in resolving key genetic mechanisms that might underlie specific clinical presentations and comorbidity patterns.^37, 38, 41–43^

In their proposal McLellan et al.^10^ note that while those with mild-to-moderate SUDs are currently not a high priority population for treatment efforts, they represent “one reasonable starting point” for defining a state similar to prediabetes, indexing accumulating risk for progression to “addiction,” and thus merit heightened vigilance and intervention. In the current study, only 27.6% of the high-risk mild-to-moderate AUD group endorsed seeking professional help or engaging in treatment. Our study provides only limited support for combining across mild and moderate AUD diagnostic categories, showing that within this broad category those endorsing certain criteria were at higher risk for severe AUD and suggesting a different categorization. Given that the hazards of progressing to severe AUD were largely equivalent between those with moderate AUD and high-risk mild-to-moderate AUD and comparatively much greater than those with mild AUD or low-risk mild-to-moderate AUD, results here indicate that mild AUD may more accurately reflect temporally-limited drinking problems consistent with endorsement of less severe criteria.^44–47^

The current study should be interpreted in light of its limitations. First, identification of high-risk criteria may be influenced by the AUD-enriched family-based study design, though comparisons to prior IRT estimates suggest results are generally consistent.^14, 15^ Second, as the longitudinal COGA cohort is primarily comprised of offspring from the cross-sectional sample, results from the two analyses can only be considered semi-independent. Third, many of the DSM-5 criteria assessed using the SSAGA are compound criteria comprised of multiple items of varying severity (e.g., social/interpersonal problems). Disaggregation of compound criteria into individual items, though not always practical in clinical settings, may help to further elucidate symptoms signaling increased risk for development of severe AUD.^48^ Fourth, while the 11 DSM-5 criteria are consistent across SUDs, findings here for AUD may not extend to other substances.

## Conclusion

We conclude that certain neurobiologically grounded high-risk criteria could improve the identification of individuals at higher risk of progression to severe AUD. Defining addiction vulnerability using criteria themselves provides greater resolution and is consistent with extant etiological and clinical insight.

## Supporting information

Supplementary Methods, Tables, and Results

## Data Availability

COGA data are available through the National Institute on Alcohol Abuse and Alcoholism or the database of Genotypes and Phenotypes (dbGaP; phs000763.v1.p1, phs000125.v1.p1). PGC alcohol dependence GWAS summary statistics may be obtained from the PGC website (https://www.med.unc.edu/pgc/). MVP GWAS summary statistics are available through dbGaP (phs001672). FinnGenR8 ICD-based AUD GWAS data were obtained from https://r8.finngen.fi/pheno/AUD. For more information, visit https://finngen.gitbook.io/documentation/.

## Acknowledgements

This research also used GWAS summary statistics data from the Psychiatric Genomics Consortium’s Substance Use Disorder working group (PGC-SUD), the Million Veterans Program (MVP), and the FinnGen Research Project Release 8 (FinnGenR8). We would like to acknowledge and thank the particpants and investigators of the many studies that made these consortia and data possible, without whom this effort would not be possible. This content is solely the responsibility of the authors and does not necessarily represent the official views of the National Institutes of Health, the Department of Veterans Affairs, or the United States Government.

The Collaborative Study on the Genetics of Alcoholism (COGA), Principal Investigators B. Porjesz, V. Hesselbrock, T. Foroud; Scientific Director, A. Agrawal; Translational Director, D. Dick, includes ten different centers: University of Connecticut (V. Hesselbrock); Indiana University (H.J. Edenberg, T. Foroud, Y. Liu, M.H. Plawecki); University of Iowa Carver College of Medicine (S. Kuperman, J. Kramer); SUNY Downstate Health Sciences University (B. Porjesz, J. Meyers, C. Kamarajan, A. Pandey); Washington University in St. Louis (L. Bierut, J. Rice, K. Bucholz, A. Agrawal); University of California at San Diego (M. Schuckit); Rutgers University (J. Tischfield, D. Dick, R. Hart, J. Salvatore); The Children’s Hospital of Philadelphia, University of Pennsylvania (L. Almasy); Icahn School of Medicine at Mount Sinai (A. Goate, P. Slesinger); and Howard University (D. Scott). Other COGA collaborators include: L. Bauer (University of Connecticut); J. Nurnberger Jr., L. Wetherill, X., Xuei, D. Lai, S. O’Connor, (Indiana University); G. Chan (University of Iowa; University of Connecticut); D.B. Chorlian, J. Zhang, P. Barr, S. Kinreich, G. Pandey (SUNY Downstate); N. Mullins (Icahn School of Medicine at Mount Sinai); A. Anokhin, S. Hartz, E. Johnson, V. McCutcheon, S. Saccone (Washington University); J. Moore, F. Aliev, Z. Pang, S. Kuo (Rutgers University); A. Merikangas (The Children’s Hospital of Philadelphia and University of Pennsylvania); H. Chin and A. Parsian are the NIAAA Staff Collaborators. We continue to be inspired by our memories of Henri Begleiter and Theodore Reich, founding PI and Co-PI of COGA, and also owe a debt of gratitude to other past organizers of COGA, including Ting-Kai Li, P. Michael Conneally, Raymond Crowe, and Wendy Reich, for their critical contributions. This national collaborative study is supported by NIH Grant U10AA008401 from the National Institute on Alcohol Abuse and Alcoholism (NIAAA) and the National Institute on Drug Abuse (NIDA).

