## Supplementary Methods, Tables, and Results for "Specific diagnostic criteria identify those at high risk for progression from ‘preaddiction’ to severe alcohol use disorder"

[1]Department of Psychiatry, Washington University School of Medicine, St. Louis, MO, USA, [2]Department of Psychiatry, Rutgers Robert Wood Johnson Medical School, Piscataway, NJ, USA, [3]Department of Psychiatry and Behavioral Sciences, State University of New York Health Sciences University, Brooklyn, NY, USA, [4]Department of Psychiatry, University of Iowa, Iowa City, IA, USA, [5]Department of Psychiatry, Indiana University, Indianapolis, IN, USA, [6]Department of Psychiatry, University of California San Diego Medical School, San Diego, CA, USA, [7]Department of Biochemistry and Molecular Biology, Indiana University, Indianapolis, IN, USA

**eMethods.** Supplemental methods
**eTable 1.** Cross-sectional COGA cohort (*N* = 13,110) item response theory severity parameters and endorsement rates for DSM-5 AUD criteria organized by diagnostic groups
**eTable 2.** All correlate sample sizes for COGA cross-sectional cohort organized by diagnostic groups
**eTable 3.** Results from mixed effect logistic AUD PGS regression models in COGA cross-sectional sample
**eTable 4**. Alcohol-related, psychiatric comorbidity, electroencephalography, and AUD polygenic score correlate sample sizes for COGA cross-sectional cohort organized by low- and high-risk mild-to-moderate and severe AUD
**eTable 5.** Results from mixed linear and logistic regression models comparing alcohol-related, psychiatric, electroencephalography, and polygenic score correlates across low-risk mild-to-moderate, high-risk mild-to-moderate, and severe AUD in COGA cross-sectional cohort
**eResults**. Supplemental survival analysis results
**eReferences**

This supplementary material has been provided by the authors to give readers additional information about their work.

**eMethods.** Supplemental methods

Assessment

Diagnostic assessment of an array of lifetime alcohol use disorder (AUD) symptoms in COGA, which can be mapped onto a variety of diagnostic classification systems, including both DSM-IV[1] and DSM-5[2], was conducted using the Semi-Structured Interview for the Genetics of Alcoholism (SSAGA), which also includes criterion-level and diagnostic assessments of other psychiatric and substance use disorders (SUDs) as well as other non-diagnostic aspects of alcohol use.[3,4] Individuals below age 18 at time of assessment were administered an adolescent SSAGA (c-SSAGA)[4,5] in both cross-sectional and longitudinal cohorts.

Alcohol Use Disorder Polygenic Scores

AUD polygenic scores (PGS) were calculated for genotyped individuals of European (EA; *n*=5,396) and African (AA; *n*=1,774) ancestry. Ancestral background was defined using both principal components analysis of array data and self-declared non-Hispanic white (EA) and non-Hispanic Black (AA) ethnicity/race. Additional details regarding genotyping, imputation, and quality control are available in a related publication.[6] Summary statistics from a genome-wide association study (GWAS) meta-analysis of data from the Million Veterans Program (MVP; $n_{case}$=45,995, $n_{control}$=221,396),[7] a "leave-one-out" GWAS of alcohol dependence from the Psychiatric Genomics Consortium (PGC) analysis that excluded the COGA sample ($n_{case}$=10,381, $n_{control}$=31,801),[8] and FinnGen Research Project Release 8 ($n_{case}$=14,307, $n_{control}$=328,192)[9] were used to code PGS using PRS-CS-auto[10] in EA individuals. PRS-CSx,[11] was used with GWAS summary statistics from both the EA analyses as well as a GWAS meta-analysis of AUD in AA individuals from PGC, not including COGA ($n_{case}$=3,335, $n_{control}$=2,945)[8] and MVP ($n_{case}$=17,267, $n_{control}$=39,381)[12] to create meta-analyzed combined weights. Individual PGSs were then calculated in PLINK 1.9[13] using PRS-CS- and PRS-CSx-derived weights. PGS were scaled to *M*=0, *SD*=1 using the *scale* function in R.

Diagnostic group (i.e., low-risk mild-to-moderate, high-risk mild-to-moderate, and severe AUD) associations with AUD PGS were examined separately for each ancestral group using mixed effect linear regression models fitted using the *lme4* package (v1.1-30)[14] controlling for age, age$^2$, sex, 10 genetic ancestry principal components, birth cohort (dummy variables representing birth years prior to 1930, 1930 to 1949, 1950 to 1969, and 1970 and after), and genotyping array type (for European subsample) as fixed effects, and for family ID as a random intercept. Mixed models were specified comparing (1) low-risk and high-risk mild-to-moderate AUD to severe AUD (reference group) and (2) low-risk mild-to-moderate and severe AUD to high-risk mild-to-moderate AUD (reference group). Additional mixed effect linear regression models with these covariates were used to examine associations between AUD PGSs and AUD criterion counts. Follow-up pairwise comparisons (e.g., high-risk *vs.* low-risk mild-to-moderate AUD, moderate *vs.* mild AUD, AUD *vs.* no AUD, etc.) were also conducted using mixed effect logistic regression models with these same covariates (**eTable 3**).

Item Response Theory Analysis

To test the assumptions of local independence and unidimensionality of one-parameter logistic item response theory (IRT) models,[15] a criteria-based confirmatory factor analysis was conducted for the full sample specifying a single AUD factor using weighted least square mean and variance adjusted estimation in the *lavaan* package (v0.6-12)[16] in R.[17] This single-factor model provided excellent fit to data (RMSEA, 0.035; 90% CI, 0.033-0.037; CFI, 0.998; TLI, 0.998; SRMR, 0.020). While two-parameter IRT models also provide information regarding the relative discrimination ability of each criteria/item, as has been suggested previously,[18,19] these models significantly complicate the goal of selecting criteria based on severity as they introduce further complexity given the additional information regarding discrimination. Thus, for the current approach, the one-parameter logistic model is preferred as it provides a more parsimonious description of the data, provides adequate fit, and is more consistent with current AUD diagnostic schemes in clinical practice (i.e., presence *vs.* absence).

Survival Analyses

The longitudinal cohort was restricted to alcohol-exposed individuals having data for at least baseline and one follow-up interview. The small number (*n*=71) of individuals who had met criteria for severe AUD at baseline were also excluded. As with the cross-sectional sample, sex, self-reported race/ethnicity, and AUD diagnostic information were measured using SSAGA/c-SSAGA interviews at each available timepoint for each individual in the longitudinal cohort (*N*=2,818). Waves of data were restructured to reflect age at each assessment (range=13-36 years). Age at onset of severe AUD was coded as the earliest assessment age at which an individual endorsed 6+ AUD criteria, while age at final assessment was used for those who did not meet severe AUD criteria during the study period (i.e., right-censored).

**eTable 1.** Cross-sectional COGA cohort ($N$ = 13,110) item response theory severity parameters and endorsement rates for DSM-5 AUD criteria organized by diagnostic groups

| | Severity Parameter *b* (SE) | Average Symptom Count *M* (*SD*) | Criteria Endorsement by Diagnostic Group (%, 95% CI) | | | | |
|---|---|---|---|---|---|---|---|
| | | | | Mild-to-Moderate AUD ("Preaddiction") | | | |
| | | | Single Criterion | Mild AUD (2-3 criteria) | Moderate AUD (4-5 criteria) | Mild-to-Moderate AUD (2-5 criteria) | Severe AUD (≥6 criteria) |
| | | | *n* = 1,649 | *n* = 2,184 | *n* = 1,295 | *n* = 3,479 | *n* = 3,298 |
| Hazardous Use | 0.07 (0.03) | 5.8 (3.3) | 37.2 [34.9, 39.6] | 64.5 [62.4, 66.5] | 80.5 [78.2, 82.5] | 70.4 [68.9, 71.9] | 93.5 [92.6, 94.3] |
| Larger/Longer | 0.14 (0.03) | 6.0 (3.2) | 23.9 [21.9, 26.0] | 63.0 [61.0, 65.1] | 85.5 [83.5, 87.4] | 71.4 [69.9, 72.9] | 96.0 [95.3, 96.6] |
| Tolerance | 0.82 (0.04) | 6.5 (3.2) | 15.8 [14.1, 17.6] | 43.9 [41.8, 46.0] | 68.5 [65.9, 71.0] | 53.1 [51.4, 54.7] | 89.7 [88.6, 90.7] |
| Cut Down | 0.91 (0.04) | 6.7 (3.1) | 15.6 [14.0, 17.5] | 34.2 [32.2, 36.2] | 70.3 [67.8, 72.8] | 47.6 [46.0, 49.3] | 91.7 [90.7, 92.6] |
| Social Interpersonal | 1.14 (0.04) | 7.2 (2.8) | 5.0 [4.1, 6.2] | 22.2 [20.5, 24.0] | 68.2 [65.6, 70.7] | 39.3 [37.7, 41.0] | 96.6 [95.9, 97.2] |
| **Failure to Fulfill** | **2.31 (0.04)** | **8.4 (2.2)** | **0.6 [0.3, 1.1]** | **4.2 [3.4, 5.1]** | **22.3 [20.1, 24.7]** | **11.0 [10.0, 12.0]** | **85.4 [84.1, 86.6]** |
| **Physical/Psychological** | **2.56 (0.04)** | **8.5 (2.3)** | **0.9 [0.5, 1.5]** | **4.7 [3.9, 5.7]** | **19.0 [17.0, 21.2]** | **10.0 [9.1, 11.1]** | **77.9 [76.5, 79.3]** |
| **Give Up** | **3.04 (0.04)** | **9.0 (1.9)** | **0.2 [0.1, 0.6]** | **1.6 [1.1, 2.2]** | **9.0 [7.5, 10.6]** | **4.3 [3.7, 5.0]** | **69.5 [67.9, 71.0]** |
| **Withdrawal** | **3.09 (0.04)** | **9.0 (1.9)** | **–** | **1.3 [0.9, 1.9]** | **9.0 [7.5, 10.6]** | **4.2 [3.6, 4.9]** | **68.3 [66.7, 69.9]** |
| **Craving** | **3.46 (0.04)** | **9.1 (2.0)** | **0.8 [0.5, 1.4]** | **1.4 [1.0, 2.0]** | **7.7 [6.4, 9.3]** | **3.8 [3.2, 4.5]** | **57.9 [56.2, 59.6]** |
| **Time Spent** | **4.21 (0.04)** | **9.4 (2.0)** | **0.1 [0.0, 0.4]** | **1.0 [0.7, 1.5]** | **5.6 [4.5, 7.0]** | **2.7 [2.2, 3.3]** | **41.0 [39.4, 42.7]** |

*Note*: AUD = alcohol use disorder; **Bold** text denotes criteria designated as high-risk based on IRT results with severity parameter values >2 (i.e., 50% endorsement probability by individuals 2+ *SD* above mean AUD latent severity). Hazardous Use = Recurrent alcohol use (3+ times) in situations in which it is physically hazardous; Larger/longer = Drinking in larger amounts or over longer periods than intended; Tolerance = Need for markedly increased amounts of alcohol to achieve intoxication or desired effect or a markedly diminished effect with continued use of the same amount of alcohol; Cut Down = Persistent desire or three or more unsuccessful efforts to stop, cut down, or control drinking; Social Interpersonal = Continued alcohol use despite having persistent or recurrent (3+ times) social or interpersonal problems caused or exacerbated by the effects of alcohol; Failure to Fulfill = Recurrent use of alcohol resulting in a failure to fulfill major role obligations at work, school, or home; Physical/Psychological = Continued drinking despite knowledge of having a persistent or recurrent physical or psychological problem that is likely to be caused or exacerbated by drinking; Give Up = Important social, occupational, or recreational activities given up or reduced because of drinking; Withdrawal = The characteristic withdrawal syndrome for alcohol; or drinking (or using a closely related substance) to relieve or avoid withdrawal symptoms; Craving = Craving, or a strong desire or urge to use alcohol; Time Spent = A great deal of time spent in activities necessary to obtain, to use, or to recover from the effects of drinking

**eTable 2.** All correlate sample sizes for COGA cross-sectional cohort organized by diagnostic groups

| Variables | No Symptoms | Single Criterion | Mild AUD (2-3 criteria) | Moderate AUD (4-5 criteria) | Mild-to-Moderate AUD | Severe AUD (≥6 criteria) |
|---|---|---|---|---|---|---|
| | *n* = 4,684 | *n* = 1,649 | *n* = 2,184 | *n* = 1,295 | *n* = 3,479 | *n* = 3,298 |
| Sociodemographic | | | | | | |
| Sex | 4,684 | 1,649 | 2,184 | 1,295 | 3,479 | 3,298 |
| Race/Ethnicity | 4,684 | 1,649 | 2,184 | 1,295 | 3,479 | 3,298 |
| Income | 4,506 | 1,598 | 2,131 | 1,267 | 3,398 | 3,236 |
| Education in Years | 4,680 | 1,649 | 2,184 | 1,294 | 3,478 | 3,297 |
| Relationship | 4,649 | 1,637 | 2,169 | 1,291 | 3,460 | 3,298 |
| Alcohol-Related | | | | | | |
| Drinking Every Day Week+ | 2,745 | 1,649 | 2,184 | 1,295 | 3,479 | 3,297 |
| No. Drinks Every Day Week+[1] | 460 | 532 | 1,157 | 958 | 2,115 | 3,085 |
| Blackouts | 2,687 | 1,649 | 2,184 | 1,295 | 3,479 | 3,297 |
| Age First Intoxication | 3,436 | 1,609 | 2,169 | 1,293 | 3,462 | 3,289 |
| Age Regular Drinking | 2,956 | 1,578 | 2,150 | 1,288 | 3,438 | 3,297 |
| Max Drinks[2] | 4,680 | 1,646 | 2,184 | 1,295 | 3,479 | 3,293 |
| Sought Help/Treatment | 2,957 | 1,647 | 2,184 | 1,295 | 3,479 | 3,298 |
| Psychiatric Comorbidity | | | | | | |
| MDD | 3,015 | 1,073 | 1,437 | 801 | 2,238 | 1,757 |
| ASPD | 4,621 | 1,614 | 2,132 | 1,237 | 3,369 | 3,017 |
| SUD | 4,684 | 1,649 | 2,184 | 1,295 | 3,479 | 3,298 |
| Theta ERO | 1,885 | 682 | 953 | 578 | 1,531 | 1,320 |
| Delta ERO | 1,885 | 682 | 953 | 578 | 1,531 | 1,320 |
| P300 Amplitude | 1,696 | 607 | 829 | 521 | 1,350 | 1,097 |
| AUD PGS | | | | | | |
| AA Subsample | 694 | 175 | 214 | 175 | 389 | 516 |
| EA Subsample | 1,671 | 688 | 1,007 | 582 | 1,589 | 1,448 |

*Note:* AUD = alcohol use disorder; MDD = major depressive disorder; ASPD = antisocial personality disorder; SUD = comorbid substance use disorder; ERO = event related oscillations; PGS = polygenic score; AA = African ancestry; EA = European ancestry

Comparison sample sizes varied across correlates according to patterns of missing data.

[1] Sample sizes for maximum number of drinks consumed every day during period of drinking every day for a week or more are restricted based on endorsement of ever drinking every day for a week or more

[2] Maximized over available interviews

**eTable 3.** Results from mixed effect logistic AUD PGS regression models in COGA cross-sectional sample

| **European ancestry subsample (*n* = 5,396)** | **OR** | **95% CI** | ***P*** |
|---|---|---|---|
| **Criterion severity** | | | |
| High-Risk Mild-to-Moderate AUD *vs.* Low-Risk Mild-to-Moderate AUD | 1.10 | [0.98, 1.24] | $1.11 \times 10^{-1}$ |
| Severe AUD *vs.* Low-Risk Mild-to-Moderate AUD | 1.08 | [0.98, 1.19] | $1.27 \times 10^{-1}$ |
| Severe AUD *vs.* High-Risk Mild-to-Moderate AUD | 0.94 | [0.84, 1.07] | $3.50 \times 10^{-1}$ |
| **DSM-5 categories** | | | |
| Moderate AUD *vs.* Mild AUD | 1.07 | [0.96, 1.19] | $2.04 \times 10^{-1}$ |
| Severe AUD *vs.* Mild AUD | 1.07 | [0.97, 1.19] | $1.76 \times 10^{-1}$ |
| Severe AUD *vs.* Moderate AUD | 0.98 | [0.88, 1.09] | $7.26 \times 10^{-1}$ |
| AUD *vs.* No AUD | **1.16** | **[1.09, 1.24]** | $\mathbf{8.69 \times 10^{-6}}$ |
| Mild-to-moderate *vs.* No AUD | **1.13** | **[1.04, 1.21]** | $\mathbf{1.97 \times 10^{-3}}$ |
| Severe AUD *vs.* No AUD | **1.23** | **[1.12, 1.35]** | $\mathbf{1.57 \times 10^{-5}}$ |

| **African ancestry subsample (*n* = 1,774)** | **OR** | **95% CI** | ***P*** |
|---|---|---|---|
| **Criterion severity** | | | |
| High-Risk Mild-to-Moderate AUD *vs.* Low-Risk Mild-to-Moderate AUD | 1.11 | [0.91, 1.35] | $3.25 \times 10^{-1}$ |
| Severe AUD *vs.* Low-Risk Mild-to-Moderate AUD | **1.30** | **[1.09, 1.56]** | $\mathbf{4.33 \times 10^{-3}}$ |
| Severe AUD *vs.* High-Risk Mild-to-Moderate AUD | 1.08 | [0.90, 1.29] | $4.13 \times 10^{-1}$ |
| **DSM-5 categories** | | | |
| Moderate AUD *vs.* Mild AUD | 1.11 | [0.91, 1.36] | $2.99 \times 10^{-1}$ |
| Severe AUD *vs.* Mild AUD | **1.27** | **[1.07, 1.51]** | $\mathbf{7.02 \times 10^{-3}}$ |
| Severe AUD *vs.* Moderate AUD | 1.10 | [0.92, 1.32] | $2.84 \times 10^{-1}$ |
| AUD *vs.* No AUD | **1.13** | **[1.01, 1.26]** | $\mathbf{3.67 \times 10^{-2}}$ |
| Mild-to-moderate *vs.* No AUD | 1.02 | [0.89, 1.17] | $7.24 \times 10^{-1}$ |
| Severe AUD *vs.* No AUD | **1.27** | **[1.10, 1.47]** | $\mathbf{1.47 \times 10^{-3}}$ |

*Note*: AUD = alcohol use disorder; PGS = polygenic score; OR = odds ratio

Fixed covariates: age, $age^2$, sex, 10 PCs, genotyping array (for European subsample), cohort

Random covariates: family ID

Ancestry grouping based on PC and reported race/ethnicity (Black non-Hispanic, white non-Hispanic)

*African ancestry sample AUD PGS on AUD symptom count: $\beta = 0.26$, SE = 0.08, $P = 1.04 \times 10^{-3}$

*European ancestry sample AUD PGS on AUD symptom count: $\beta = 0.20$, SE = 0.05, $P = 9.36 \times 10^{-6}$

**eTable 4**. Alcohol-related, psychiatric comorbidity, electroencephalography, and AUD polygenic score correlate sample sizes for COGA cross-sectional cohort organized by low- and high-risk mild-to-moderate and severe AUD

| | Mild-to-Moderate AUD ("Preaddiction") | | |
|---|---|---|---|
| | Endorsed Low-Risk Criteria | Endorsed High-Risk Criteria | Severe AUD |
| Variables | *n* = 2,486 | *n* = 993 | *n* = 3,298 |
| Alcohol-Related | | | |
| Drinking Every Day Week+ | 2,486 | 993 | 3,297 |
| No. Drinks Every Day Week+[1] | 1,422 | 693 | 3,085 |
| Blackouts | 2,486 | 993 | 3,297 |
| Age First Intoxication | 2,474 | 988 | 3,289 |
| Age Regular Drinking | 2,459 | 979 | 3,297 |
| Max Drinks[2] | 2,486 | 993 | 3,293 |
| Sought Help/Treatment | 2,486 | 993 | 3,298 |
| Psychiatric Comorbidity | | | |
| MDD | 1,627 | 611 | 1,757 |
| ASPD | 2,437 | 932 | 3,017 |
| SUD | 2,486 | 993 | 3,298 |
| Theta ERO | 1,094 | 437 | 1,320 |
| Delta ERO | 1,094 | 437 | 1,320 |
| P300 Amplitude | 966 | 384 | 1,097 |
| AUD PGS | | | |
| AA Subsample | 204 | 185 | 516 |
| EA Subsample | 1,204 | 385 | 1,448 |

*Note:* AUD = alcohol use disorder; MDD = major depressive disorder; ASPD = antisocial personality disorder; SUD = comorbid substance use disorder; ERO = event related oscillations; PGS = polygenic score; AA = African ancestry; EA = European Ancestry Comparison sample sizes varied across correlates according to patterns of missing data.

[1] Sample sizes for maximum number of drinks consumed every day during period of drinking every day for a week or more are restricted based on endorsement of ever drinking every day for a week or more

[2] Maximized over available interviews

**eTable 5.** Results from mixed linear and logistic regression models comparing alcohol-related, psychiatric, electroencephalography, and polygenic score correlates across low-risk mild-to-moderate, high-risk mild-to-moderate, and severe AUD in COGA cross-sectional cohort

| | High-risk mild-to-moderate AUD *vs.* low-risk mild-to-moderate AUD | | Severe AUD *vs.* high-risk mild-to-moderate AUD | | Severe AUD *vs.* low-risk mild-to-moderate AUD | |
|---|---|---|---|---|---|---|
| Variables | OR/β | 95% CI | OR/β | 95% CI | OR/β | 95% CI |
| Alcohol-Related | | | | | | |
| Drinking Every Day Week+ | 1.14 | [0.93, 1.39] | **4.04** | **[3.12, 5.23]** | **4.61** | **[3.45, 6.15]** |
| No. Drinks Every Day Week+ | **0.15** | **[0.05, 0.25]** | **0.52** | **[0.42, 0.61]** | **0.67** | **[0.55, 0.78]** |
| Blackouts | 0.96 | [0.79, 1.15] | **2.55** | **[2.06, 3.15]** | **2.44** | **[1.91, 3.11]** |
| Age First Intoxication | 0.08 | [0.00, 0.15] | **-0.29** | **[-0.37, -0.21]** | **-0.21** | **[-0.31, -0.11]** |
| Age Regular Drinking | 0.06 | [-0.02, 0.13] | **-0.26** | **[-0.34, -0.18]** | **-0.20** | **[-0.30, -0.11]** |
| Max Drinks | **0.08** | **[0.01, 0.15]** | **0.58** | **[0.50, 0.66]** | **0.66** | **[0.57, 0.75]** |
| Sought Help/Treatment | **1.48** | **[1.19, 1.85]** | **7.18** | **[5.86, 8.81]** | **10.63** | **[8.24, 13.72]** |
| Psychiatric Comorbidity | | | | | | |
| MDD | **1.53** | **[1.12, 2.10]** | **2.07** | **[1.45, 2.94]** | **3.17** | **[2.07, 4.83]** |
| ASPD | **1.56** | **[1.17, 2.08]** | **1.84** | **[1.40, 2.42]** | **2.87** | **[2.04, 4.04]** |
| SUD | **1.34** | **[1.09, 1.63]** | **2.61** | **[2.08, 3.28]** | **3.49** | **[2.70, 4.52]** |
| Theta ERO | **-0.12** | **[-0.24, 0.00]** | -0.09 | [-0.22, 0.04] | **-0.21** | **[-0.36, -0.06]** |
| Delta ERO | 0.00 | [-0.12, 0.12] | -0.05 | [-0.18, 0.08] | -0.05 | [-0.20, 0.10] |
| P300 Amplitude | -0.08 | [-0.20, 0.03] | -0.10 | [-0.22, 0.03] | **-0.18** | **[-0.32, -0.04]** |
| AUD PGS | | | | | | |
| EA Subsample | 0.09 | [-0.01, 0.20] | 0.02 | [-0.09, 0.13] | 0.07 | [-0.01, 0.16] |
| AA Subsample | 0.12 | [-0.28, 0.05] | 0.12 | [-0.05, 0.28] | **0.22** | **[0.06, 0.38]** |

*Note:* AUD = alcohol use disorder; MDD = major depressive disorder; ASPD = antisocial personality disorder; SUD = comorbid substance use disorder; ERO = event related oscillations; PGS = polygenic score; AA = African ancestry; EA = European ancestry; OR = odds ratio; β = standardized regression coefficient

**eResults**. Supplemental results from survival analyses

In the longitudinal sample (*N*=2,818), a total of 252 individuals eventually met criteria for severe AUD (i.e., 8.9% of total sample; median onset age of 24 years). Cox proportional hazards regression models demonstrated increasing hazards as a function of prior criterion count-based severity: single (*n*=566) *vs.* no criteria (*n*=1,236; adjusted hazard ratio [aH], 1.13; 95% CI, 0.66-1.93); mild AUD (*n*=833; aHR, 3.48; 95% CI, 2.37-5.11; 14.7% transition); and moderate AUD (*n*=183; aHR, 11.30; 95% CI, 7.33-17.43; 40.4% transition; between mild and moderate AUD aHR, 3.25; 95% CI, 2.10-5.03). In total, 1,016 individuals in the sample (36.1%) met criteria for mild-to-moderate AUD prior their final assessment, and 196 of these individuals (19.3%) went on to develop severe AUD at a subsequent timepoint. Cox proportion hazards analyses similarly found that a prior mild-to-moderate AUD diagnosis was associated with a significantly elevated hazards of severe AUD after adjusting for covariates (aHR, 11.30; 95% CI, 7.33-17.43; Figure 2), though this result was primarily driven by those endorsing high-risk criteria (see main text).

Other youth characteristics that have been previously linked to progression to severe AUD were also significantly associated with progression to severe AUD; however, adjusted hazards for these characteristics ranged from aHR, 0.75-0.79 for age at onset variables (i.e., first drink, regular drinking, and first intoxication; inverse aHR, 1.27-1.33) to aHR=7.88 for other SUDs, suggesting in general that these indices are not as predictive of transitioning to severe AUD as either a prior moderate AUD diagnosis or a high-risk mild-to-moderate AUD diagnosis. In multivariate models including all risk factor variables in addition to covariates mentioned above (i.e., accounting for shared variance among these feature), high-risk mild-to-moderate was the strongest predictor (aHR, 4.25; 95% CI, 2.57-7.04) followed by other SUDs (aHR, 3.65; 95% CI, 2.42-5.52). Similar models for prior mild *vs.* moderate AUD also demonstrated an association between a prior diagnosis of moderate AUD and progression to severe AUD (aHR, 4.46; 95% CI, 2.70-7.35) that was larger in magnitude than other youth characteristics (other SUDs aHR, 3.76; 95% CI, 2.49-5.68).
